# Customize Deep Learning-based De-Identification Systems Using Local Clinical Notes - A Study of Sample Size

**DOI:** 10.1101/2020.08.09.20171231

**Authors:** Xi Yang, Jiang Bian, Yonghui Wu

**Affiliations:** Health Outcomes and Biomedical Informatics, College of Medicine, University of Florida, Gainesville FL USA

**Keywords:** Clinical notes de-identification, Deep learning, Sample size

## Abstract

Electronic Health Records (EHRs) are a valuable resource for both clinical and translational research. However, much detailed patient information is embedded in clinical narratives, including a large number of patients’ identifiable information. De-identification of clinical notes is a critical technology to protect the privacy and confidentiality of patients. Previous studies presented many automated de-identification systems to capture and remove protected health information from clinical text. However, most of them were tested only in one institute setting where training and test data were from the same institution. Directly adapting these systems without customization could lead to a dramatic performance drop. Recent studies have shown that fine-tuning is a promising method to customize deep learning-based NLP systems across different institutes. However, it’s still not clear how much local data is required. In this study, we examined the customizing of a deep learning-based de-identification system using different sizes of local notes from UF Health. Our results showed that the fine-tuning could significantly improve the model performance even on a small local dataset. Yet, when the local data exceeded a threshold (e.g., 700 notes in this study), the performance improvement became marginal.

## 1 INTRODUCTION

Rapid adoption of Electronic Health Records (EHRs) systems has made EHRs data an important resource for clinical and translational research [1,2]. The Clinical narratives are a unique type of data summarizing the detailed communications between patients and health providers in free text format. Many important clinical information, such as family history, social determinants of health, and drug adverse events [3–6], can only be extracted from clinical text. Meanwhile, clinical text contains various personal private information of patients, such as their names, date of birth, address, and social security numbers, which is associated with a high risk of privacy breaching [7]. Under the Health Insurance Portability and Accountability Act (HIPAA) Privacy Rule, all the identifiable health information should be secured and protected for only medical purposes [8,9]. The HIPAA common rule emphasizes that, even for clinical research, it is required to either obtain consent from patients or a waiver from an Institutional Review Board (IRB) [10] to use health data containing protected health information (PHI). De-identification [11–13] is a key technology to remove these PHIs from clinical text to facilitate researchers using clinical notes with minimum risk of violating the HIPPA private rules in their studies.

The HIPAA Safe Harbor method defines 18 categories of PHI to be removed from health data to generate de-identified clinical data. The de-identification systems for clinical notes usually have two fundamental steps including (1) detecting PHIs in free text and (2) substituting them with predefined replacements. Manually de-identification is time-consuming and expensive given the large volume of text data. Therefore, researchers have explored natural language processing (NLP) for automating the de-identification procedure of clinical text. The clinical NLP community has organized several shared tasks [12–14] to solicit state-of-the-art systems for clinical notes de-identification and contributed several publicly available datasets [17,18] as benchmarks to facilitate de-identification research. Many deep learning-based models [11,19–22] demonstrated promising performances during these challenges. However, most of these models were evaluated using training and test data from the same institute with similar note types (i.e., one institute setting) [23].

Recently, several studies have shown that directly adopting these challenge winning models for local clinical corpora de-identification without customization could lead to dramatic performance drop [24,25], as training and test data are from different institutes. There is a cross institute issue when applying the state-of-the-art deep learning models for de-identification. Several studies [25,26] have shown that the fine-tuning strategy was a promising customization approach to enhance the deidentification performances of deep learning-based models in cross institute settings. However, the efficiency of the fine-tuning method was not comprehensively assessed previously. Questions as “how many annotated notes are required for fine-tuning” and “how to decide whether model performances are saturated” have not been answered.

In this study, we systematically examined methods to customize deep learning-based de-identification systems trained using the open challenge dataset and local corpora with a various number of clinical notes. We assessed the performances of using different sized local clinical text to customize various deep learning-based models through fine-tuning. This study identified the reasonable number of local clinical text required to customize deep learning-based de-identification systems.

## 2 MATERAIL AND METHOD

### 2.1 Dataset

In this study, we collected a total number of 1,100 clinical notes distributed in 39 different note types (e.g., progress, H&P, and Radiology Report) from the UF Health Integrated Data Repository (IDR). Annotators manually annotated PHIs in these notes. We randomly divided the annotated notes into a training set of 900 notes and a test set of 200 notes stratified by the note types. We used the training set to develop de-identification models and use the test set for evaluation. To assess performances of using various sizes (denoted as N) of local text, we experimented with five different sizes of local notes, including 100, 300, 500, 700, and 900 (i.e., all of the local notes) notes, respectively. For each size, the notes were randomly selected from the whole training set (N=900) with the same random seed. For each training set, we further split it into a short training set and a validation set with a size ratio of 9:1. The description of the datasets was summarized in Table 1.

**Table 1:** Description of the datasets for de-identification.

| Dataset | Number of Notes |  |  |
| --- | --- | --- | --- |
|  | train | dev | test |
| UF Health | 810 | 90 | 200 |
| subset N=100 | 90 | 10 | 200 |
| subset N=300 | 270 | 30 | 200 |
| subset N=500 | 450 | 50 | 200 |
| subset N=700 | 630 | 70 | 200 |

### 2.2 Models and Training Strategies

In this study, we explored a deep learning-based de-identification model - the Long-short Term Memory – Conditional Random Fields (LSTM-CRFs). We trained the models using the training datasets and test their performances using the test set. We compared two training strategies, including the fine-tuning and the training-from-scratch. For the fine-tuning approach, the deep learning model was first pre-trained using a de-identification dataset curated in the 2014 i2b2 challenge [25] as a base checkpoint. Then, we continuously fine-tuned this checkpoint (i.e., initialize new models with the weights from this checkpoint and use the same model settings) using the local UF datasets (i.e., different number of notes) developed in this study. For the strategy of training-from-scratch, we did not adopt any pre-trained models and trained new models from scratch on each training set. For comparison, we used the 2014 i2b2 pre-trained model (N=0) as a baseline.

### 2.3 PHI categories

Although the HIPAA Safe Harbor method defines 18 PHI categories for de-identification, directly using these definitions is infeasible in practice, and customization is required. For example, the “Geographic information smaller than state” contains various types of location-related concepts, and all the identifiable numbers (e.g., Medical Record Number, Account Number, Social Security Number) can be treated merely as ID. In addition, the PHIs of face photos and biometric identifiers (e.g., fingerprints) are rarely presented in the clinical notes. Therefore, we followed the 2014 i2b2 challenge and remapped the 18 categories of the Safe Harbor PHIs to a new set of 13 PHI types including person names, age, date, phone (for phone and fax), web (for internet-related information like URL, email), ID, institute names, zip code, PO Box, street name, city, location other (for location-related information but cannot be categories), and other (for all other information that can be used for re-identification). Among all the newly defined PHIs, the name (NAME), date (DATE), ID, and institute name (INSTITUTE) contain the information with a high risk of re-identification [27]. Therefore, we paid particular attention to the four PHI categories.

### 2.4 Experiments and Evaluation

We adopted the LSTM-CRFs model developed in our previous works [25,28] using TensorFlow [29]. We trained models using the short-training sets and selected the optimized model checkpoints according to the performances on the validation sets. We adopted a pre-trained word embeddings contained two million-word vectors developed using the fastText algorithm on the Common Crawl corpus [30]. We set the following parameters for the LSTM-CRFs model: the word embedding dimension was 300; the character embedding dimension was 25; the bidirectional word-level LSTM had an output dimension of 100; the bidirectional character-level LSTM had an output size of 25; the learning rate was fixed at 0.005; the input layer for the word-level LSTM applied a dropout at a probability of 0.5; the stochastic gradient descending used a gradient clapping at [− 5.0, 5.0] and a momentum term fixed at 0.9. For the fine-tuning strategy, we set the training epochs to 20, while the training epochs used for the training-from-scratch method was set to 30. For evaluation, we calculated the model level performance as micro-averaged strict precision, recall, and F1-score. We also reported the F1-scores for the PHIs of DATE, NAME, ID, and INSTITUTE achieved by the fine-tuning models.

## 3 RESULTS

Figure 1 compares the performance of using two training strategies with a various number of local notes. The baseline i2b2 model without customization (the green point in Figure 1 at N=0) only achieved an F1-score of 0.8186 on the UF Health test set. After customizing using 100 UF notes (N=100), the fine-tuned model achieved an F1-score of 0.9181 while the performance of the training-from-scratch (trained using UF data only) model was 0.8900. Compared to the baseline, the fine-tuned model remarkably improved the performance by ∼10% and outperformed the training-from-scratch model by ∼3%. The performances achieved by the fine-tuned models were consistently better than the training-from-scratch models across all different sized training sets (blues vs. reds in Figure 1). For N=300, 500, 700, and 900, the fine-tuned model achieved the F1 scores of 0.9443, 0.9622, 0.9707, and 0.9734, respectively. While the performances obtained by the models only trained with the UF dataset were 0.9367, 0.9542, 0.9676, and 0.9681 under the same experiment settings. Compared to the best training-from-scratch model (red at N=900 in Figure 1; F1-score of 0.9681), the fine-tuned model trained with 700 notes (blue at N=700 in Figure 1) already achieved a better performance (0.9707 for fine-tuning vs. 0.9681 for training-from-scratch).

**Figure 1:**
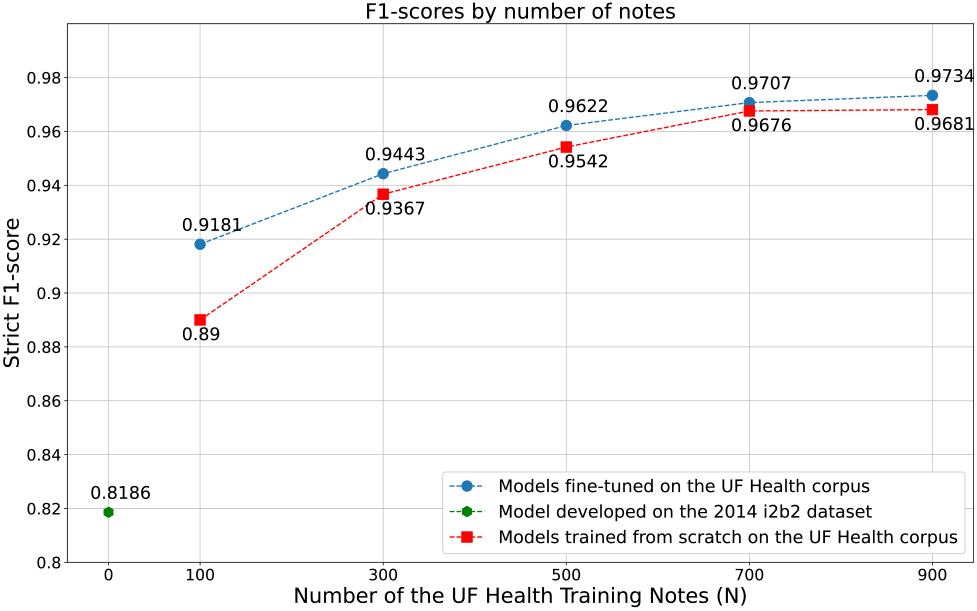
The micro-averaged performances by the different sizes of training sets.

Figure 2 plotted the F1-scores for PHIs of DATE, NAME, ID, and INSTITUTE against the sizes of the training sets. For the baseline i2b2 model without fine-tuning (N=0), the F1-scores for NAME, ID, and INSTITUTE were remarkably lower than 0.9 as 0.8007, 0.5529, and 0.1737, while the F1-score for DATE was 0.948. Fine-tuning the model with only 100 local notes could significantly improve the performances of DATE, NAME, ID, and INSTITUTE to 0.9741, 0.9195, 0.7395, and 0.6842, respectively. Training with more local notes could continuously help models to achieve better performance for each PHI category. However, such improvements became less significant when the training set size was over 500 notes (N > 500). With N=500, the fine-tuned model already achieved the performances over 0.9 for all four PHIs categories. Compared to the fine-tuned model customized using 900 local notes, the model customized with 700 local notes obtained a comparable performance for all four PHI categories (0.9856 vs. 0.9871 for DATE; 0.9694 vs. 0.9725 for NAME; 0.9616 vs. 0.9543 for ID; 0.9327 vs. 0.9406 for INSTITUTE). Among all 13 types of PHIs, the “street names” and “location other” were challenges for de-identification models to detect. For the model fine-tuned with 900 notes, the F1-scores of city names and location other were only 0.8007 and 0.8800, which were significantly lower compared to other PHIs.

**Figure 2:**
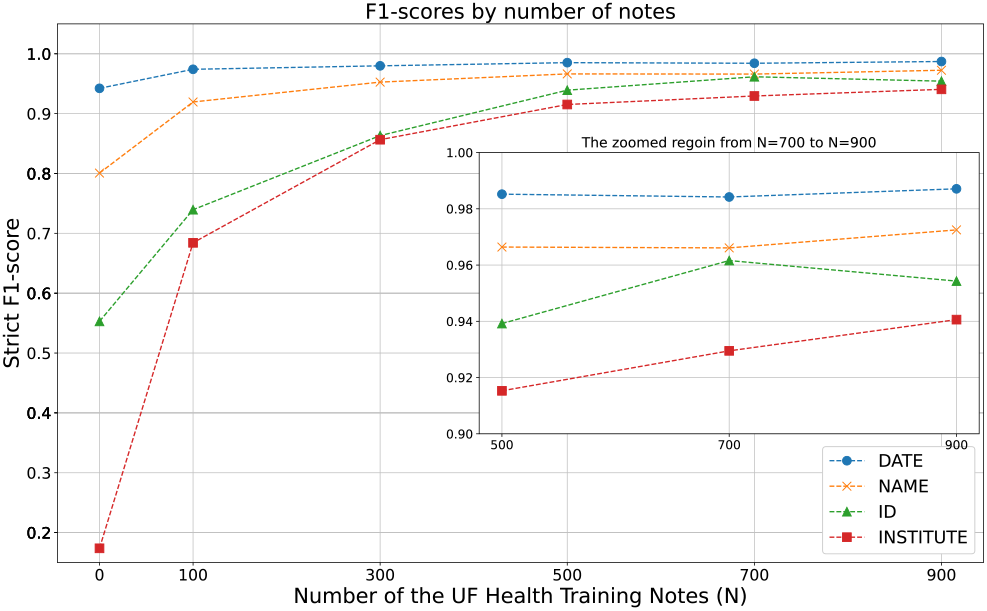
The fine-tuning model performances on the PHI categories of DATE, NAME, ID, and INSTITUTE across different training sets.

## 4 DISCUSSION AND CONCLUSION

Annotating local notes is essential for adapting deep learning-based de-identification models in cross institute settings. In this study, we explored the sample size issue when customizing these deep learning models using local data. We examined the model performance using different sizes of local annotated notes for customizing. Our results showed that the fine-tuning strategy is a better approach compared with the training-from-scratch method for de-identification of clinical text. Models developed via fine-tuning consistently yielded better performances across all training datasets with a various number of notes from 100 to 900. In addition, the fine-tuning models required significantly less training data to achieve similar or even better performances. Since manually annotating PHIs in clinical notes is often expensive and time-consuming, the fine-tuning strategy can speed up the development of de-identification systems and save costs.

Although the results (Figure 1) illustrated that customizing with more clinical notes could continuously enhance the performance of detecting PHIs, the improvements became marginal when the training set size exceeds 500 notes. Especially for the PHI categories of NAME, DATE, and ID, the model fine-tuned with 700 local notes already obtained decent performances (F1-scores ∼ 0.95). Extra training notes (e.g., N=900) did not significantly contribute to further model performance improvement, especially for PHIs like names, IDs, and dates.

This study demonstrated that customizing existing deep learning models developed using public datasets using local data is an efficient method for de-identification of clinical text.

## Data Availability

Data will not be available to public

## ACKNOWLEDGMENTS

We would like to thank the i2b2 challenge organizers to provide the annotated corpus. We gratefully acknowledge the support of NVIDIA Corporation with the donation of the GPUs used for this research.

This study was partially supported by a Patient-Centered Outcomes Research Institute® (PCORI®) Award (ME-2018C3-14754), a grant from the National Cancer Institute, 1R01CA246418 R01, a grant from the National Institute on Aging, NIA R21AG062884, the University of Florida (UF) Informatics Institute Junior SEED Program (00129436), and the Cancer Informatics and eHealth core jointly supported by the UF Health Cancer Center and the UF Clinical and Translational Science Institute. The content is solely the responsibility of the authors and does not necessarily represent the official views of the funding institutions.

